# Classification of Adolescent Drinking via Behavioral, Biological, and Environmental Features: A Machine Learning Approach with Bias Control

**DOI:** 10.64898/2026.02.24.26347002

**Authors:** Ruobing Liu, Mohamed Azzam, Nikki Zabik, Shibiao Wan, Jennifer Blackford, Jieqiong Wang

**Affiliations:** Department of Neurological Sciences, University of Nebraska Medical, Omaha, NE 68198, USA; Munroe-Meyer Institute for Genetics and Rehabilitation, University of Nebraska Medical Center, Omaha, NE 68106, USA; Department of Genetics, Cell Biology and Anatomy, University of Nebraska Medical Center, Omaha, NE 68198, USA

**Keywords:** Adolescent alcohol use, alcohol drinking classification, machine learning, brain measures

## Abstract

In 2024, approximately 30% of U.S. adolescents reported having consumed alcohol at least once in their lifetime, with about 25% of these individuals engaging in binge drinking. Adolescent alcohol use is associated with neurodevelopmental impairments, elevated risk of later alcohol use, and mental health disorders. These findings underscore the importance of identifying the variables driving adolescent alcohol use and leveraging them for early identification and targeted intervention. Previous studies have typically developed machine-learning classification models that use neuroimaging data in combination with limited clinical measurements. Neuroimaging data are expensive and difficult to obtain at scale, whereas clinical measures are more practical for large-scale screening due to their low cost and widespread accessibility. However, clinical-only approaches for alcohol drinking classification remain largely underexplored. Furthermore, prior studies have often focused on adults, limiting generalizability to the broader adolescent population. Additionally, confounding factors such as age and substance use, which are strongly correlated with alcohol consumption, have often been inadequately addressed, potentially inflating classification performance. Finally, class imbalance remains a persistent challenge, with prior attempts yielding only limited improvements. To address these limitations, we propose *FocalTab*, a framework that integrates TabPFN with focal loss for robust generalization and effective mitigation of class imbalance. The approach also incorporates an initial preprocessing step to remove confounding factors to account for age and substance-use. We compare *FocalTab* against state-of-the-art methods across different variable selections and dataset settings. *FocalTab* achieves the highest accuracy (84.3%) and specificity (80.0%) in the most stringent setting, in which both age and substance use variables were excluded, whereas competing models drop to near-chance specificity (12-24%). We further applied SHapley Additive exPlanations (SHAP) analysis to identify key clinical predictors of drinker classification, supporting enhanced screening and early intervention.

## 1. Introduction

Underage drinking refers to alcohol drinking among youth under 21 years of age. According to recent reports from the Substance Abuse and Mental Health Services Administration ^1^, over 30% of U.S. adolescents have consumed alcohol at least once, and approximately 25% of these individuals engage in binge drinking ^2^. Adolescent binge drinking is linked to both short- and long-term adverse outcomes, including poor academic performance, increased accident risk, mental health disorders, violence, liver complications, and a higher risk of future alcohol use disorder (AUD) ^3,4^. These consequences also impose substantial societal costs, including increased healthcare spending, public safety burdens, and productivity losses. Therefore, developing accurate classification models to identify adolescent drinkers’ initiation and understanding the key variables that drive this transition is critical for enabling early identification and targeted intervention, which can significantly reduce the long-term risk of alcohol dependence and related health consequences ^5–7^.

While machine learning (ML) methods have been increasingly applied to classify alcohol use in adults ^8–10^, their application to adolescent populations remains underexplored. Among adolescent-focused studies, most have relied on neuroimaging data and have only focused on a specific, relatively short age range. Whelan et al. s^11^ (2014) applied an elastic net to multimodal data, including structural Magnetic Resonance Imaging (MRI), task-based fMRI (functional Magnetic Resonance Imaging), personality, cognitive, environmental, and genetic measures, from 692 adolescents aged 14 years in the IMAGEN study to classify current binge drinkers from non-binge drinkers, achieving an accuracy of 91%. Ottino-González et al. ^12^ (2024) used structural covariance networks (SCN) derived from FreeSurfer cortical thickness measures alongside an ML classifier on data from 272 adolescents aged 17-22 in the NCANDA study to classify heavy alcohol users from matched controls, demonstrating robust and generalizable classification across cohorts. In practice, these imaging data are often inaccessible for many adolescents, as high acquisition costs and the requirement to live near an MRI facility impose significant financial and geographical barriers, restricting the diversity and representativeness of study samples. In contrast, clinical measurements (demographics, interview data, etc.) provide a more accessible and scalable alternative for studying alcohol consumption, enabling larger and more inclusive cohorts, which is essential for achieving higher accuracy and better performance.

Many prior studies on adolescent drinker detection ^13–17^ have not explicitly addressed age bias, leaving their models vulnerable to age-driven classification. Age represents a major source of bias ^18,19^, as alcohol use increases with age. In addition, several clinical measures are themselves correlated with age, imposing an indirect source of bias. For example, studies in adolescents ^20–22^ have shown that increasing age is associated with continued improvement in working memory capacity. Steinberg et al. (2008) ^23^ reported that both impulsivity and the effectiveness of parental monitoring decrease with increasing age among adolescents ^24^. Laursen & Veenstra (2021) ^25^ noted that peer conformity peaks during early adolescence. Consequently, models trained on age-related inputs may inadvertently learn age-dependent patterns rather than alcohol-specific signals.

Many studies ^13,26,27^ used different types of substance use (cigarettes, cannabis, etc.) as classification variables for alcohol drinking classification, ignoring substance use bias. These variables are strongly associated with alcohol use, where alcohol often represents an early initiating substance that precedes later drug experimentation ^28^. Incorporating these variables as predictors may artificially inflate model performance without capturing the underlying behavioral or psychosocial risk factors that independently contribute to alcohol misuse. Furthermore, many adolescents are not exposed to cigarettes or other substances ^29^, resulting in substantial class imbalance. This trend is supported by national data showing that, among adolescents aged 12 to 17, the prevalence of cigarette use declined to 2.3% in 2019 ^30^, the prevalence of e-cigarette use declined to around 5% in 2020 ^31–35^, and the prevalence of marijuana use also declined significantly in 2021 ^36^. In addition, even when adolescents did use substances, self-report measures were subject to significant underreporting due to social desirability bias, particularly for sensitive and stigmatized behaviors ^37–39^.

Class imbalance represents another major challenge in adolescent alcohol-drinking classification, as datasets typically contain substantially more non-drinkers than drinkers ^1,2^. To address this issue, prior studies have employed strategies such as SMOTE-based oversampling ^17,40^, majority-class undersampling ^16,39^, or extensive cross-validation procedures ^13^. However, these methods have inherent limitations: 1) SMOTE may generate synthetic samples that do not accurately reflect the minority-class distribution, potentially introducing noise and overfitting; 2) undersampling discards informative majority-class observations, reducing statistical power; and 3) repeated cross-validation does not fundamentally correct the imbalance during model training. In contrast, focal loss ^42^ offers a more principled alternative by down-weighting the contribution of majority-class samples and emphasizing minority-class examples during training, thereby enabling more effective use of the available data and improving classification performance under imbalanced settings.

As illustrated in Fig 1, our proposed work addresses these methodological gaps through several key contributions. First, we develop a classification framework to detect adolescent drinking utilizing exclusively clinical measurements, enhancing accessibility and scalability for real-world screening applications. Second, we expand our subjects’ age range from 12 to 22 years, encompassing early adolescence through young adulthood, thereby including substantially more subjects while capturing the full trajectory of adolescent neurodevelopment. Third, we employ confound regression to remove age-related variance from predictor variables, addressing the developmental biases that have limited prior work. Fourth, we construct a methodologically rigorous learning system that explicitly excludes substance use variables from the feature set, thereby eliminating potential data leakage and the confounding relations between alcohol and substance use and reducing the dependence on substance use. Finally, we incorporate focal loss to address the inherent class imbalance in adolescent drinking data, preserving maximum sample utilization while avoiding the distributional distortions introduced by synthetic oversampling. Collectively, these methodological innovations yield a reliable, generalizable classification system for distinguishing drinkers from non-drinkers that addresses the key limitations of existing approaches in this field.

**Figure 1.**
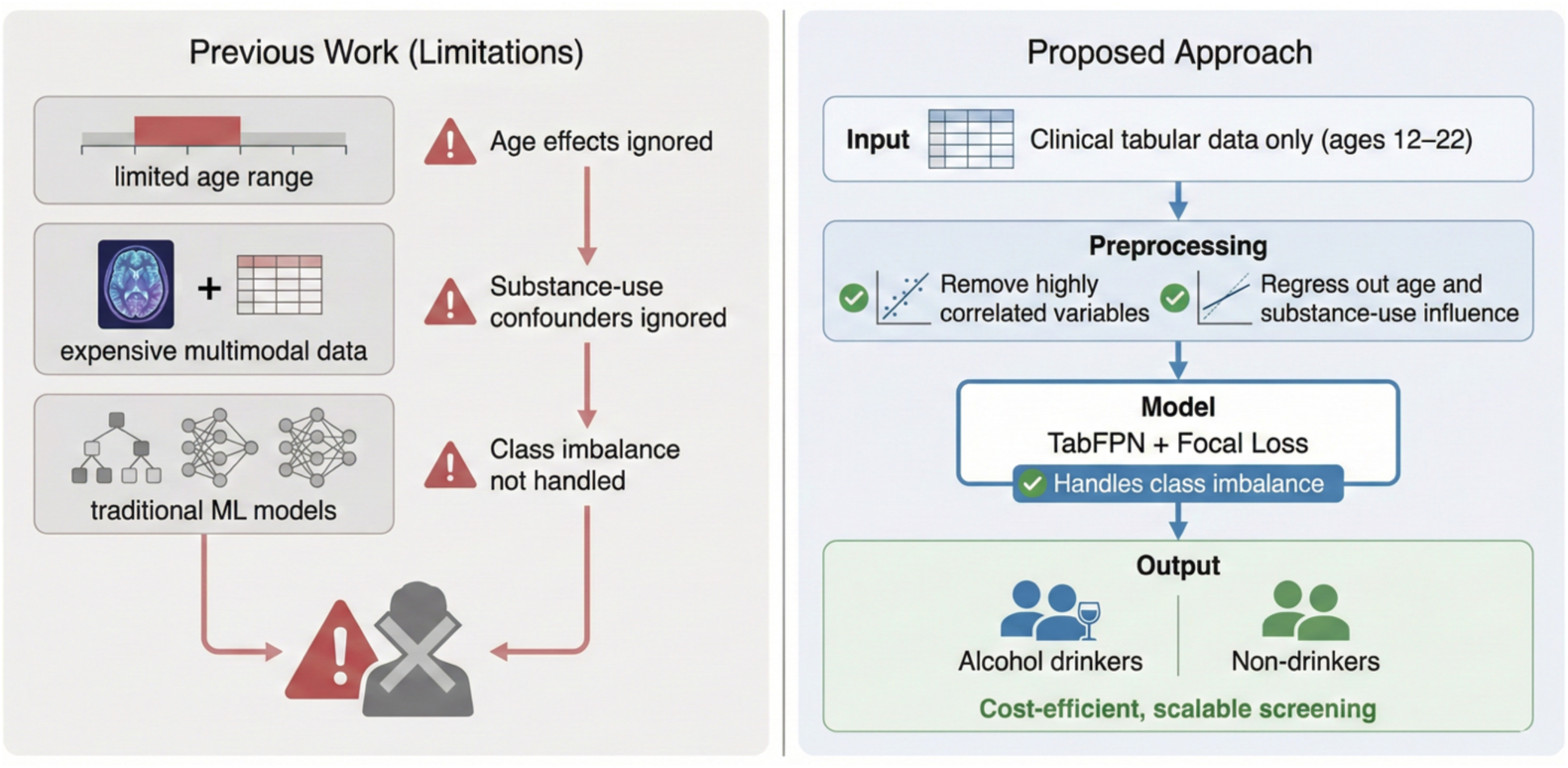
Comparison between prior adolescent alcohol-use detection approaches and the proposed framework.

## 2. Materials and Methods

### 2.1. Participants

This study used data from the National Consortium on Alcohol and Neurodevelopment in Adolescence (NCANDA), a multisite longitudinal study designed to examine the effects of alcohol exposure on adolescent neurodevelopment ^43^. Using an accelerated longitudinal design, NCANDA initially enrolled a sample of 831 adolescents aged 12 to 21 and conducted annual follow-up assessments. The baseline data were analyzed in this study to classify non-/lower drinkers (shortened as non-drinkers) versus moderate/heavy drinkers (shortened as drinkers) status using machine learning approaches.

The definitions of non-drinkers and drinkers followed the modified Cahalan inventory ^44^ using responses from the Customary Drinking and Drug Use Record ^45^. Non-drinkers met the following criteria: (1) drinking frequency of less than once per month or less than once per year; (2) consumption of no more than three drinks per month; and (3) no binge drinking episodes. Drinkers encompassed both moderate and heavy drinking categories. Moderate drinkers were defined as those reporting: (1) low frequency with high quantity (e.g., less than once per month with three to four drinks on average and four to five drinks maximum); or (2) moderate frequency with low quantity (e.g., once per week with two drinks on average and less than four drinks maximum). Heavy drinkers were defined as those reporting: (1) moderate frequency with high quantity (e.g., twice per month with three to four drinks on average and more than four drinks maximum); or (2) high frequency with moderate consumption (e.g., once or more per week with one to four drinks on average and four or fewer drinks maximum). There were 9 participants without baseline data, and 21 participants without any follow-up data, excluded from analyses. Based on these definitions, the final 801 samples included 661 non-drinkers and 140 drinkers.

### 2.2. Feature Selection

To ensure the reliability and independence of input features for machine learning, we applied the following feature selection criteria: 1) removal of constant features that showed no variability across participants; 2) exclusion of features with >50% missing values; and 3) elimination of features directly related to substance use, to prevent confounding or circularity in predicting drinking behavior. After applying these criteria, a total of 167 baseline features across behavioral, biological, and environmental domains were retained for model development. Table 1 lists these 167 variables across 13 categories. Alcohol expectancy (7 variables) assessed beliefs about alcohol effects using the Alcohol Expectancy Questionnaire (AEQ) ^46^. Family history (4 variables) captured familial substance use disorders via the Family History Assessment Module (FHAM) ^47^. Socioeconomic status (4 variables) included parental education, family income, and composite SES scores ^48^. Personality traits (5 variables) were measured using the Ten-Item Personality Inventory (TIPI) ^49^ and the UPPS-P Impulsive Behavior Scale (UPPS-P) ^50^. Environmental atmosphere (48 variables) encompassed neighborhood characteristics, parental monitoring, peer influences, and school climate. Child behavior and daily functioning (21 variables) assessed sleep patterns, executive function, and coping strategies using the Behavior Rating Inventory of Executive Function (BRIEF) ^51^ and Responses to Stress Questionnaire (RSQ) ^52^. Medical history (5 variables) included traumatic brain injury and hospitalizations. Demographics (5 variables) captured family structure and educational engagement. Physical measures (3 variables) included height, weight, and BMI. Pubertal development (2 variables) was assessed using the Pubertal Development Scale (PDS) ^53^. Peer relationships (1 variable) measured social support quality via the Social Support Questionnaire (SSQ) ^54^. Daily routine (31 variables) captured activities and competence. Psychiatric symptomatology (31 variables) assessed depression, conduct disorder, attention-deficit/hyperactivity disorder (ADHD), obsessive-compulsive disorder (OCD), posttraumatic stress disorder (PTSD), panic, and anxiety using the Semi-Structured Assessment for the Genetics of Alcoholism (SSAGA) ^55^ and Center for Epidemiologic Studies-Depression Scale (CES-D) ^56^.

**Table 1.**
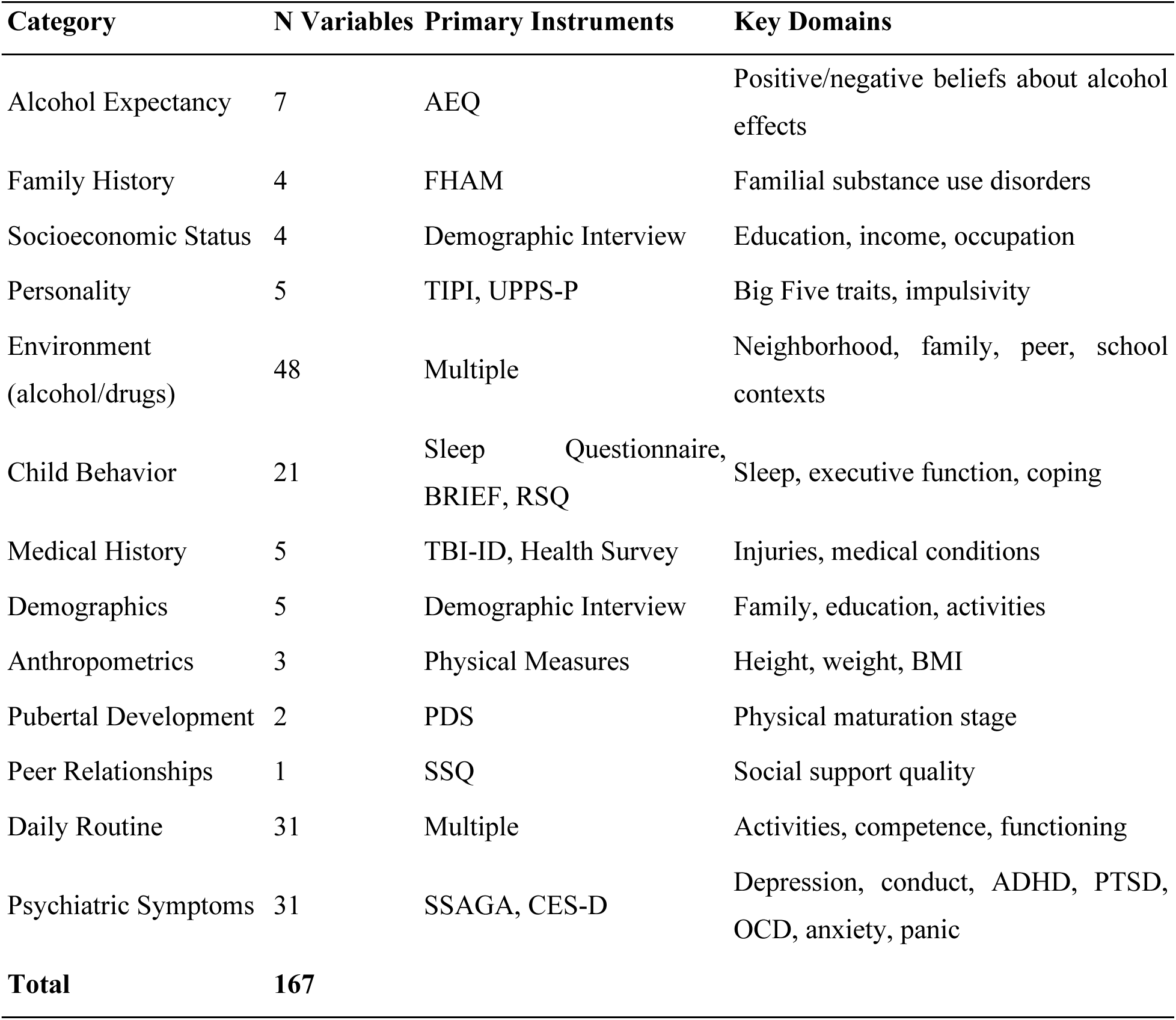
Variable Categories and Measurement Instruments.

| Category | N Variables | Primary Instruments | Key Domains |
| --- | --- | --- | --- |
| Alcohol Expectancy | 7 | AEQ | Positive/negative beliefs about alcohol effects |
| Family History | 4 | FHAM | Familial substance use disorders |
| Socioeconomic Status | 4 | Demographic Interview | Education, income, occupation |
| Personality | 5 | TIPI, UPPS-P | Big Five traits, impulsivity |
| Environment<br>(alcohol/drugs) | 48 | Multiple | Neighborhood, family, peer, school contexts |
| Child Behavior | 21 | Sleep Questionnaire,<br>BRIEF, RSQ | Sleep, executive function, coping |
| Medical History | 5 | TBI-ID, Health Survey | Injuries, medical conditions |
| Demographics | 5 | Demographic Interview | Family, education, activities |
| Anthropometrics | 3 | Physical Measures | Height, weight, BMI |
| Pubertal Development | 2 | PDS | Physical maturation stage |
| Peer Relationships | 1 | SSQ | Social support quality |
| Daily Routine | 31 | Multiple | Activities, competence, functioning |
| Psychiatric Symptoms | 31 | SSAGA, CES-D | Depression, conduct, ADHD, PTSD, OCD, anxiety, panic |
| **Total** | **167** |  |  |

### 2.3. Classification of Drinkers

We developed a machine learning model to classify adolescent drinkers versus non-drinkers using baseline clinical data from the NCANDA dataset. The overall workflow is summarized in Figure 2, which illustrates the data preprocessing strategies, model development, and evaluation procedures. Briefly, multiple input feature configurations were constructed to explicitly address potential confounding effects of age and substance use variables, followed by strategies to handle class imbalance. The processed data were then used to train and evaluate classification models. Among the evaluated models, we implemented TabPFN (Tabular Prior-Data Fitted Network) ^57^ with focal loss ^42^ (*FocalTab*) as the primary model, given its strong performance on tabular clinical data and its ability to improve minority-class learning. The detailed methodology is described in the following subsections: Section 2.3.1 outlines the model development pipeline, including input feature configurations, data preprocessing, and the TabPFN architecture with focal loss; Section describes the training strategy, including dataset balancing and cross-validation; and Section presents the evaluation framework, including comparisons with other classification models, approaches for handling class imbalance, and strategies for mitigating age- and substance use-related bias. Post-hoc analyses were subsequently conducted to enhance model interpretability and biological relevance, with feature importance analysis described in Section 2.3.4.

**Figure 2.**
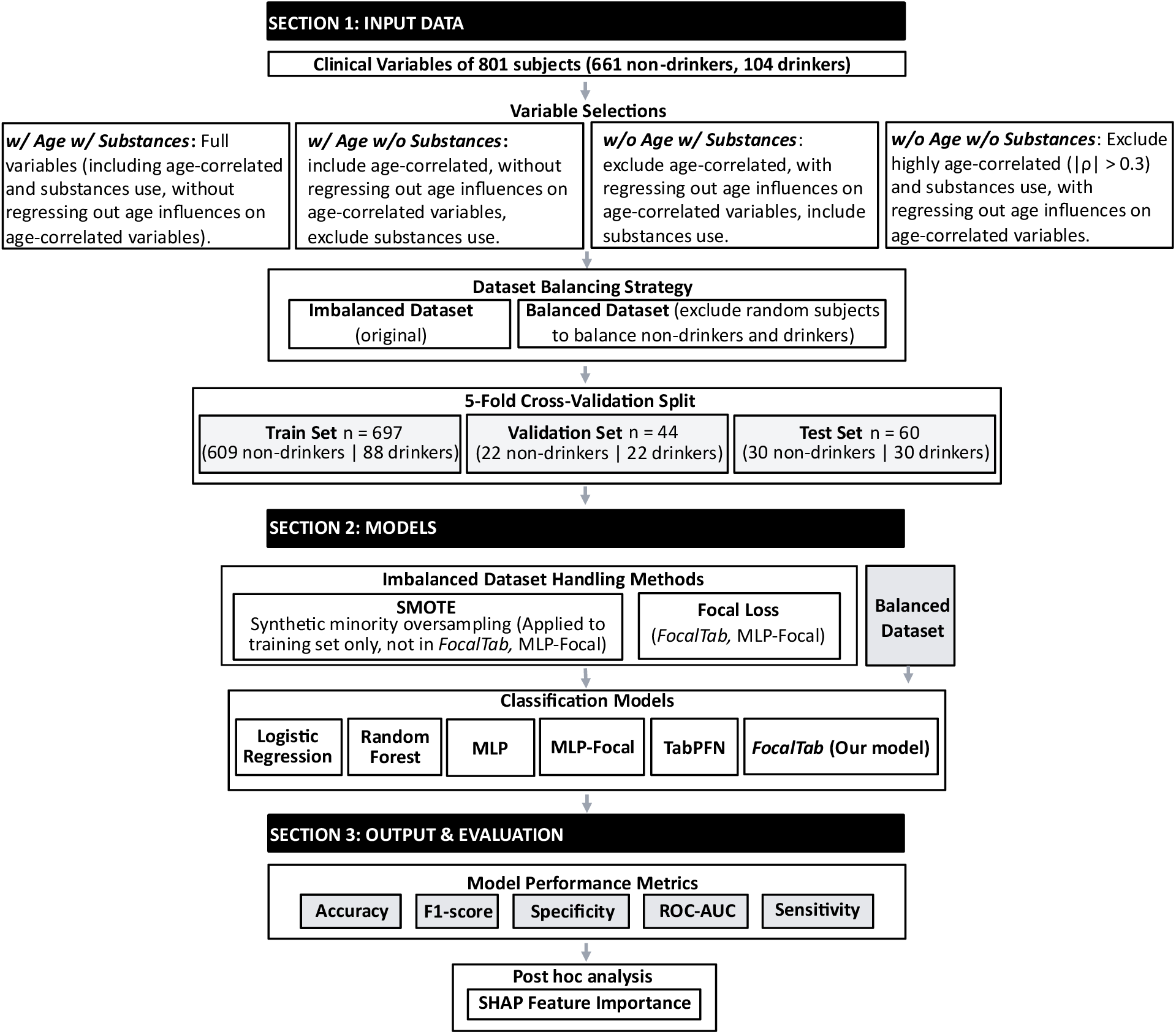
Pipeline of data preprocessing, model development, and evaluation procedures.

#### 2.3.1. Model Development via *FocalTab*

In the preprocessing step, we excluded substance use-related variables, regressed out age effects, and removed variables highly correlated with age to mitigate age-related bias (see Section 2.3.3 for details). The resulting feature set was then used as input to the *FocalTab* model to classify participants as drinkers versus non-drinkers.

TabPFN ^57^ is a pretrained, transformer-based classification foundation model designed for tabular classification via in-context learning. Unlike conventional machine learning methods that require iterative training on each new dataset, TabPFN leverages knowledge acquired during an offline pretraining phase on large collections of synthetic datasets. The architecture uses alternating row and column attention mechanisms and approximates Bayesian inference in a single forward pass.

The model accepts training samples D_train_ = {(x_1_, y_1_), …, (x_n_, y_n_)} and test variables x_test_ as set-valued inputs, yielding the posterior predictive distribution (PPD) q_θ_ (y |x_test_, D_train_) without requiring gradient-based optimization on the target dataset ^57^. The PPD for a test sample x_test_is approximated as:

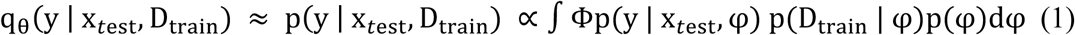

where φ represents hypotheses from a prior space Φ that includes structural causal models (SCMs) and Bayesian neural networks (BNNs), weighted by their likelihood given the training data and prior probability ^57^.

To address the substantial class imbalance in our dataset (661 non-drinkers vs. 140 drinkers), we employed focal loss ^42^ as the model’s loss function, which down-weights the contribution of easily classified examples and focuses training on difficult, misclassified samples. The focal loss is defined as:

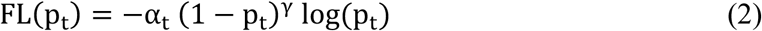

where α_t_ and γ are the parameters to be optimized, while p_t_ represents the model’s estimated probability for the ground-truth class:

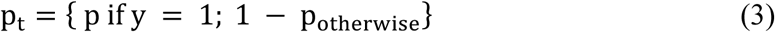

#### 2.3.2. Training Strategy

The TabPFN model was optimized using a multi-staged approach designed to maximize predictive performance while rigorously segregating training and evaluation data. Initially, we conducted an extensive grid search to identify optimal hyperparameters, fine-tuning the model on the training set across a spectrum of learning rates (5×10^−^⁶ to 1×10^−^²) and optimizers (RAdam, Adam, SGD, and RMSprop). To address potential class imbalance, Focal Loss was employed, with hyperparameters γ and α tuned via incremental intervals on the validation set. The optimal classification threshold and hyperparameter configuration were determined based on the maximum F1-score and AUC achieved during validation. Subsequently, the model was retrained on the combined training and validation cohorts using the previously identified optimal settings to ensure robust feature representation. Finally, the performance of the refitted model was assessed on a strictly held-out test set, with all classification thresholds fixed from the validation phase to ensure an unbiased evaluation of the model’s generalizability.

#### 2.3.3. Evaluation

We evaluated our model across three dimensions: (1) comparison of model performance with other state-of-the-art methods using 5-fold cross-validation (CV), (2) assessment of the impact of class imbalance strategies, and (3) examination of substance use and age biases in classifying adolescent drinkers from non-drinkers.

To further assess the performance of our model, we compared it with other methods, including (1) logistic regression ^58^, (2) random forest ^59^, (3) multilayer perceptron (MLP) ^60^, and (4) TabPFN without focal loss. For the MLP, we implemented two variants: MLP with default binary cross-entropy loss (MLP) ^61^ and MLP with focal loss (MLP-Focal). Model performance was evaluated using 5-fold CV. In each fold, subjects were randomly divided into training, validation, and test sets. The training set contained 697 subjects (609 non-drinkers and 88 drinkers), the validation set contained 44 subjects (22 non-drinkers and 22 drinkers), and the test set contained 60 subjects (30 non-drinkers and 30 drinkers). Average accuracy, F1-score, specificity, and AUC were calculated across the 5-fold CV results to evaluate each model.

To evaluate the impact of class imbalance on model performance, we compared four sampling strategies: (1) *Imbalanced-Original*, using the original class distribution; (2) Focal loss, applying focal loss to *Imbalanced-Original*; (3) *Imbalanced-SMOTE*, using the Synthetic Minority Over-sampling Technique 30 to generate synthetic drinkers samples equal to the non-drinkers class size; and (4) *Balanced*, randomly downsampling non-drinkers subjects in the training set to match the drinkers count (n = 88 per group). Validation and test sets were kept identical across strategies to ensure comparability.

To evaluate the impact of substance use variables on classification of drinkers versus non-drinkers, we compared four variable selection strategies: (1) *w/ Age w/ Substances*, all variables included, comprising substance use variables and highly age-correlated variables, without regressing out age effects; (2) *w/o Age w/o Substances*, substance use variables and highly age-correlated variables excluded, with age effects regressed out from age-correlated variables; (3) *w/ Age w/o Substances*, substance use variables excluded while highly age-correlated variables retained, without regressing out age effects; and (4) *w/o Age w/ Substances*, substance use variables retained while highly age-correlated variables excluded, with age effects regressed out from age-correlated variables.

To mitigate age-related confounding effects on classification performance, we implemented a multi-stage variable filtering strategy. Variables with strong age correlations (|ρ| > 0.3; FDR-corrected) were excluded. Variables with moderate but significant correlations (q < 0.05 after FDR correction) were residualized using linear regression: numeric variables via ordinary least squares, *x*_residual_ = *x* − (*β*_0_ + *β*_1_ ⋅ age), and binary variables via logistic regression residuals, *x*_residual_ = *x* − *p̂*(*x* = 1 ∣ age). The age variable was then removed from the feature matrix. Remaining numeric variables were standardized using parameters derived from the training set only.

#### 2.3.4. Model Interpretation with SHAP

SHapley Additive exPlanations (SHAP) ^62^ is a widely used framework for interpreting machine learning models by quantifying the contribution of individual variables to model prediction. In this study, we applied SHAP to interpret the *FocalTab* model and identify the most influential variables for drinkers’ classification. Specifically, SHAP values were computed for all input variables to estimate their marginal contributions to the predicted drinking outcomes. Variables were subsequently ranked according to their mean absolute SHAP values, enabling identification of the most important predictors driving model performance.

## 3. Results

### 3.1. Demographics Analysis

As shown in Table 2, drinker subjects (18.55 ± 1.98 years) were significantly older than non-drinker subjects (15.65 ± 2.31 years; p < 0.001). There is no significant difference of gender distribution between groups (p = 0.200). Racial composition was similar across groups, with European-American participants comprising the majority in both the non-drinkers (70.8%) and drinkers (76.4%) groups; no significant differences were observed in any racial category among each group (all p > 0.1).

**Table 2.** Demographic Characteristics of non-drinkers and drinkers Groups.

| Variable | Non-drinkers | Drinkers | p value |
| --- | --- | --- | --- |
| N | 661 | 140 |  |
| Age (in years) | 15.65 ± 2.31 | 18.55 ± 1.98 | < 0.001 *** |
| **Gender** |  |  |  |
| Female | 326 (49.3%) | 78 (55.7%) | 0.199 |
| Male | 335 (50.7%) | 62 (44.3%) |  |
| **Race** |  |  |  |
| Native American | 3 (0.5%) | 0 (0.0%) | 0.578 |
| Asian | 50 (7.6%) | 11 (7.9%) |  |
| Pacific Islander | 3 (0.5%) | 1 (0.7%) |  |
| African-American | 78 (11.8%) | 14 (10.0%) |  |
| European-American | 468 (70.8%) | 107 (76.4%) |  |
| Other | 59 (8.9%) | 7 (5.0%) |  |

*Note:* Values are presented as mean ± standard deviation for continuous variables (age) and count (percentage) for categorical variables (gender and race). Statistical comparisons between groups were conducted using independent-samples t-tests for age and chi-square tests for categorical variables of gender and race. Significance levels: \* $p < 0.05$ , \*\* $p < 0.01$ , \*\*\* $p < 0.001$ .

### 3.2. Comparison of Classification Performance with Different Models

#### 3.2.1. Model Comparison

Table 3 and Figure 3 present comparisons of six models’ performance for classifying drinkers versus non-drinkers using the *Imbalanced-Original* dataset with the *w/o Age w/o Substances* variable setting. *FocalTab* demonstrated the most robust performance under strict confound control, achieving an AUC of 0.902, accuracy of 0.843, F1-score of 0.850, and sensitivity and specificity of 0.800 (Table 3). Notably, the specificity of all other models was substantially lower than that of *FocalTab* under these settings.

**Figure 3.**
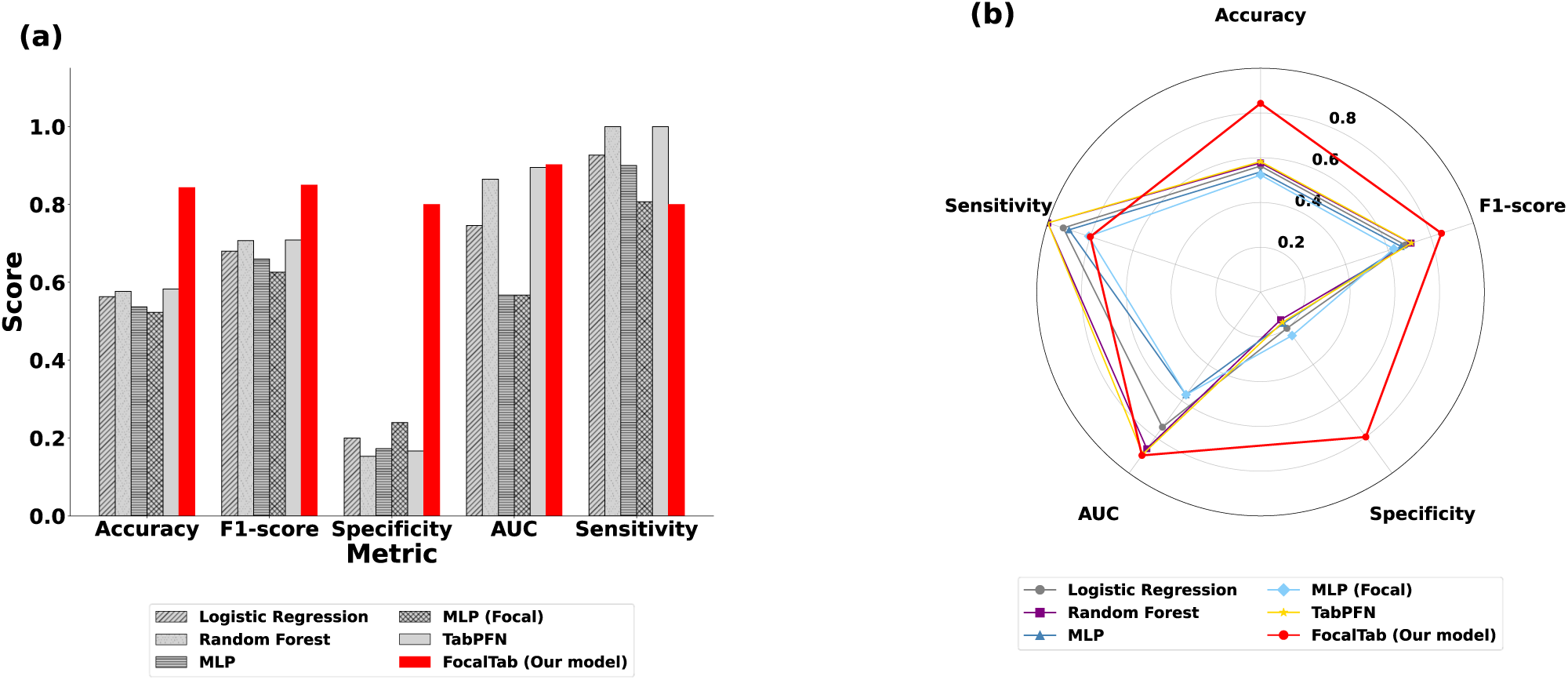
Model performance comparison for drinkers classification using the *Imbalanced-Original* dataset with the *w/o Age w/o Substances* variable setting. **(a).** Bar plot of performance metrics across models; *FocalTab* is highlighted in red. **(b).** Radar plot of performance metrics across models; *FocalTab* is highlighted in red. MLP: multilayer perceptron. AUC: ROC-AUC.

**Table 3.** Comparison of model performance for drinkers classification using the *Imbalanced-Original* dataset with the *w/o Age w/o Substances* variable setting. MLP: multilayer perceptron. AUC: ROC-AUC. Bold: *FocalTab*.

| Metric | Logistic Regression | Random Forest | MLP | MLP (Focal) | TabPFN | *FocalTab* (Our model) |
| --- | --- | --- | --- | --- | --- | --- |
| Accuracy | 0.563 | 0.577 | 0.537 | 0.523 | 0.583 | **0.843** |
| F1-score | 0.680 | 0.707 | 0.660 | 0.626 | 0.709 | **0.850** |
| Specificity | 0.200 | 0.153 | 0.173 | 0.240 | 0.167 | **0.800** |
| AUC | 0.746 | 0.865 | 0.567 | 0.567 | 0.895 | **0.902** |
| Sensitivity | 0.927 | 1.000 | 0.9000 | 0.8067 | 1.000 | **0.800** |

#### 3.2.2. Substance Use and Age Biases

Table 4 and Figures 4-5 summarize the performance of six models in classifying drinkers versus non-drinkers under the *Imbalanced-Original* dataset, highlighting the effects of substance use and age biases. Across models, performance generally decreased as biases were removed, indicating that many classifiers relied on age-related developmental variance and/or substance use variables to achieve high accuracy.

**Figure 4.**
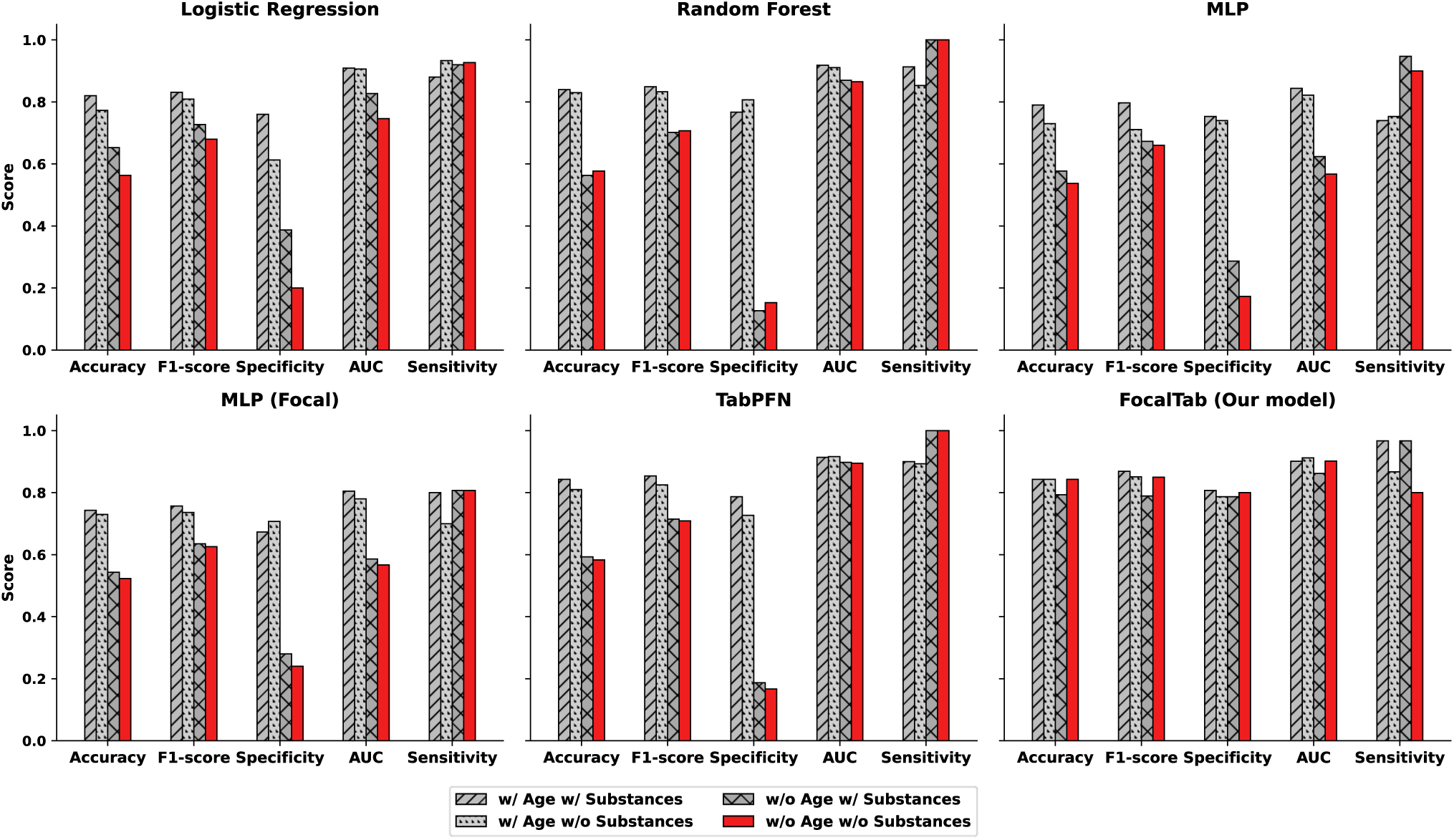
Bar plots of model performance across variable selection strategies for evaluating substance use and age biases using the Imbalanced-Original dataset setting. From left to right, top to bottom: Logistic Regression, Random Forest, MLP, MLP-Focal, TabPFN, and *FocalTab*. MLP: multilayer perceptron. AUC: ROC-AUC.

**Figure 5.**
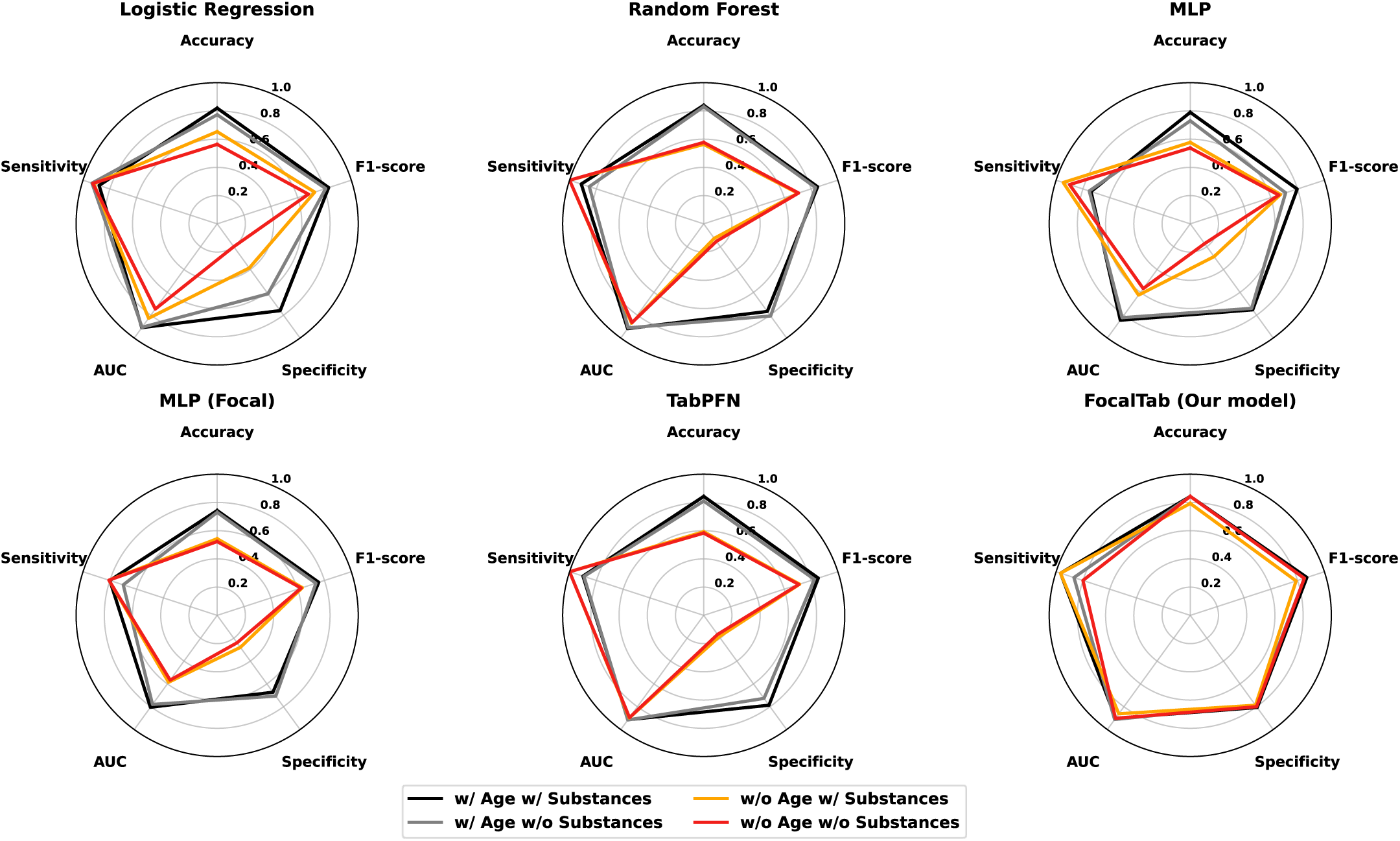
Radar plots of model performance across variable selection strategies for evaluating substance use and age biases using the *Imbalanced-Original* dataset setting. From left to right, top to bottom: Logistic Regression, Random Forest, MLP, MLP-Focal, TabPFN, and *FocalTab*. MLP: multilayer perceptron. AUC: ROC-AUC.

**Table 4.** Comparison of model performance across variable selection strategies for evaluating substance use and age biases using the *Imbalanced-Original* dataset setting. MLP: multilayer perceptron. AUC: ROC-AUC. Bold: *FocalTab*.

| Dataset Setting | Metric | Logistic Regression | Random Forest | MLP | MLP (Focal) | TabPFN | *FocalTab* (Our model) |
| --- | --- | --- | --- | --- | --- | --- | --- |
| *w/ Age w/ Substances* | Accuracy | 0.820 | 0.840 | 0.790 | 0.743 | 0.843 | **0.843** |
|  | F1-score | 0.831 | 0.849 | 0.797 | 0.757 | 0.854 | **0.869** |
|  | Specificity | 0.760 | 0.767 | 0.753 | 0.673 | 0.787 | **0.807** |
|  | AUC | 0.909 | 0.918 | 0.844 | 0.805 | 0.914 | **0.901** |
|  | Sensitivity | 0.880 | 0.913 | 0.740 | 0.800 | 0.900 | **0.967** |
| *w/ Age w/o Substances* | Accuracy | 0.773 | 0.830 | 0.730 | 0.730 | 0.810 | **0.843** |
|  | F1-score | 0.809 | 0.833 | 0.711 | 0.736 | 0.825 | **0.851** |
|  | Specificity | 0.613 | 0.807 | 0.740 | 0.707 | 0.727 | **0.787** |
|  | AUC | 0.906 | 0.911 | 0.822 | 0.780 | 0.916 | **0.912** |
|  | Sensitivity | 0.933 | 0.853 | 0.753 | 0.700 | 0.893 | **0.867** |
| *w/o Age w/ Substances* | Accuracy | 0.653 | 0.563 | 0.577 | 0.543 | 0.593 | **0.793** |
|  | F1-score | 0.727 | 0.702 | 0.673 | 0.635 | 0.715 | **0.789** |
|  | Specificity | 0.387 | 0.127 | 0.287 | 0.280 | 0.187 | **0.787** |
|  | AUC | 0.827 | 0.870 | 0.624 | 0.586 | 0.898 | **0.862** |
|  | Sensitivity | 0.920 | 1.000 | 0.947 | 0.807 | 1.000 | **0.967** |
| *w/o Age w/o Substances* | Accuracy | 0.563 | 0.577 | 0.537 | 0.523 | 0.583 | **0.843** |
|  | F1-score | 0.680 | 0.707 | 0.660 | 0.626 | 0.709 | **0.850** |
|  | Specificity | 0.200 | 0.153 | 0.173 | 0.240 | 0.167 | **0.800** |
|  | AUC | 0.746 | 0.865 | 0.567 | 0.567 | 0.895 | **0.902** |
|  | Sensitivity | 0.927 | 1.000 | 0.900 | 0.807 | 1.000 | **0.800** |

Figure 4 illustrates a consistent performance ordering across the four variable conditions: *w/ Age w/ Substances* (highest) > *w/ Age w/o Substances* > *w/o Age w/ Substances* > *w/o Age w/o Substances* (lowest). The decline was most pronounced for specificity, suggesting that removing the age and substance use confounds primarily reduced the models’ ability to correctly identify non-drinkers. Specifically, specificity fell sharply from the *w/ Age w/ Substances* to *w/o Age w/o Substances* conditions for logistic regression (0.760 to 0.200), random forest (0.767 to 0.153), MLP (0.753 to 0.173), and TabPFN (0.787 to 0.167; Table 4). This pattern indicates that, once age and substance use biases were controlled, conventional models misclassified non-drinkers at substantially higher rates.

As shown in Table 4, *w/ Age w/o Substances* (retaining age-correlated variables without age regression) typically outperformed *w/o Age w/ Substances* (retaining substance-use variables while regressing out age effects), suggesting that developmental variance contributes more strongly to model discrimination than substance-use variables in this dataset. For example, Random Forest achieved AUC = 0.919 under *w/ Age w/o Substances*, but dropped to AUC = 0.878 under *w/o Age w/ Substances,* using *Imbalanced-Original* setting (Table 4). Similar trends were observed across other models, consistent with age-linked features serving as a major driver of classification separability.

#### 3.2.3. Class Imbalance

Table 5 and Figures 6-7 present comparisons of six models’ performance for classifying drinkers versus non-drinkers using the *w/o Age w/o Substances* variable setting, revealing the effects of class imbalance on classification. Model performance under the *Balanced* setting was slightly better than under the two *Imbalanced* settings. Under the *Imbalanced-SMOTE* and *Balanced* settings, where *FocalTab* was not evaluated, TabPFN achieved the highest AUCs (0.879 with *Imbalanced-SMOTE*; 0.912 with *Balanced*); however, specificity remained substantially lower (0.107 and 0.540, respectively) than *FocalTab’s* specificity of 0.800 on the *Imbalanced-Original* setting (Table 5). Notably, *FocalTab* demonstrated the most robust performance in the *Imbalanced-Original* setting, achieving an AUC of 0.902, an accuracy of 0.843, an F1-score of 0.850, and sensitivity and specificity of 0.800 (Table 5). This specificity substantially exceeded that of all competing methods under the same *w/o Age w/o Substances* condition, indicating that *FocalTab* can correctly identify non-drinkers even on highly imbalanced datasets.

**Figure 6.**
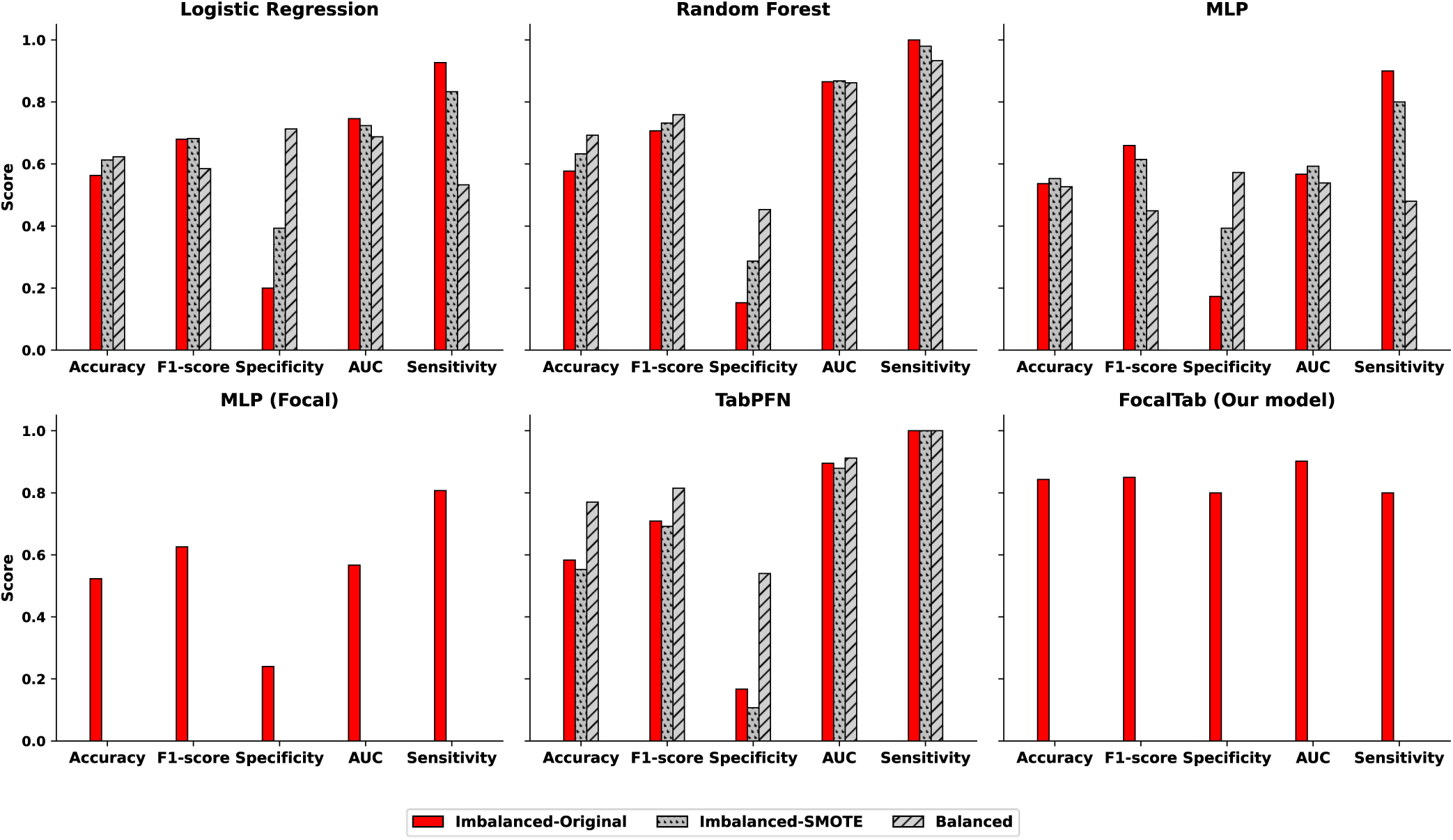
Bar plots of model performance across class imbalance strategies using the *Exclude* variable setting. From left to right, top to bottom: Logistic Regression, Random Forest, MLP, MLP-Focal, TabPFN, and *FocalTab*. MLP: multilayer perceptron. AUC: ROC-AUC.

**Figure 7.**
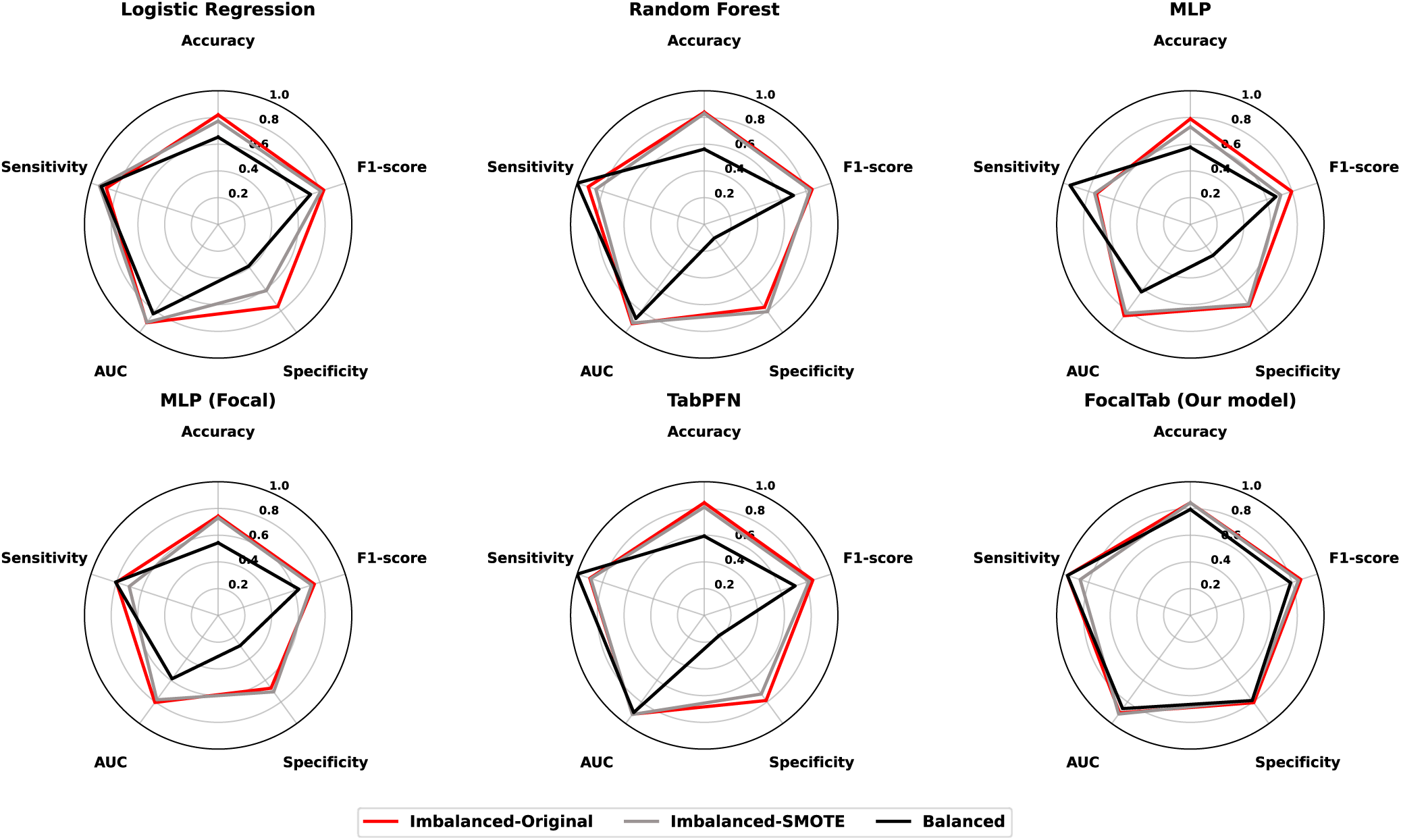
Radar plots of model performance across class imbalance strategies using the w/o Age w/o Substances variable setting. From left to right, top to bottom: Logistic Regression, Random Forest, MLP, MLP-Focal, TabPFN, and *FocalTab.* MLP: multilayer perceptron. AUC: ROC-AUC.

**Table 5.** Comparison of model performance across class imbalance strategies using the *w/o Age w/o Substances* variable setting. MLP: multilayer perceptron. AUC: ROC-AUC. Bold: *FocalTab*.

| Dataset Setting | Metric | Logistic Regression | Random Forest | MLP | MLP (Focal) | TabPFN | *FocalTab* (Our model) |
| --- | --- | --- | --- | --- | --- | --- | --- |
| *Imbalanced-Original* | Accuracy | 0.563 | 0.577 | 0.537 | 0.523 | 0.583 | **0.843** |
|  | F1-score | 0.680 | 0.707 | 0.660 | 0.626 | 0.709 | **0.850** |
|  | Specificity | 0.200 | 0.153 | 0.173 | 0.240 | 0.167 | **0.800** |
|  | AUC | 0.746 | 0.865 | 0.567 | 0.567 | 0.895 | **0.902** |
|  | Sensitivity | 0.927 | 1.000 | 0.900 | 0.807 | 1.000 | **0.800** |
| *Imbalanced-SMOTE* | Accuracy | 0.613 | 0.633 | 0.553 | — | 0.553 | — |
|  | F1-score | 0.682 | 0.732 | 0.615 | — | 0.692 | — |
|  | Specificity | 0.393 | 0.287 | 0.393 | — | 0.107 | — |
|  | AUC | 0.724 | 0.868 | 0.593 | — | 0.879 | — |
|  | Sensitivity | 0.833 | 0.980 | 0.800 | — | 1.000 | — |
| *Balanced* | Accuracy | 0.623 | 0.693 | 0.527 | — | 0.770 | — |
|  | F1-score | 0.585 | 0.759 | 0.449 | — | 0.815 | — |
|  | Specificity | 0.713 | 0.453 | 0.573 | — | 0.540 | — |
|  | AUC | 0.688 | 0.862 | 0.539 | — | 0.912 | — |
|  | Sensitivity | 0.533 | 0.933 | 0.480 | — | 1.000 | — |

### 3.3. Model Interpretation with SHAP

Figure 8 presents the top 10 most important variables with the highest mean absolute SHAP values from the *FocalTab* model for drinkers classification using the *Imbalanced-Original* dataset with the *w/o Age w/o Substances* variable setting. Higher values indicate greater variable importance. The most influential variables included alcohol expectancy (anticipated change of social behavior, sexual enhancement, improved cognitive and motor ability), psychiatric symptoms (panic, OCD, PTSD), sleep schedule, making friends, place to go at night, and how to spend money.

**Figure 8.**
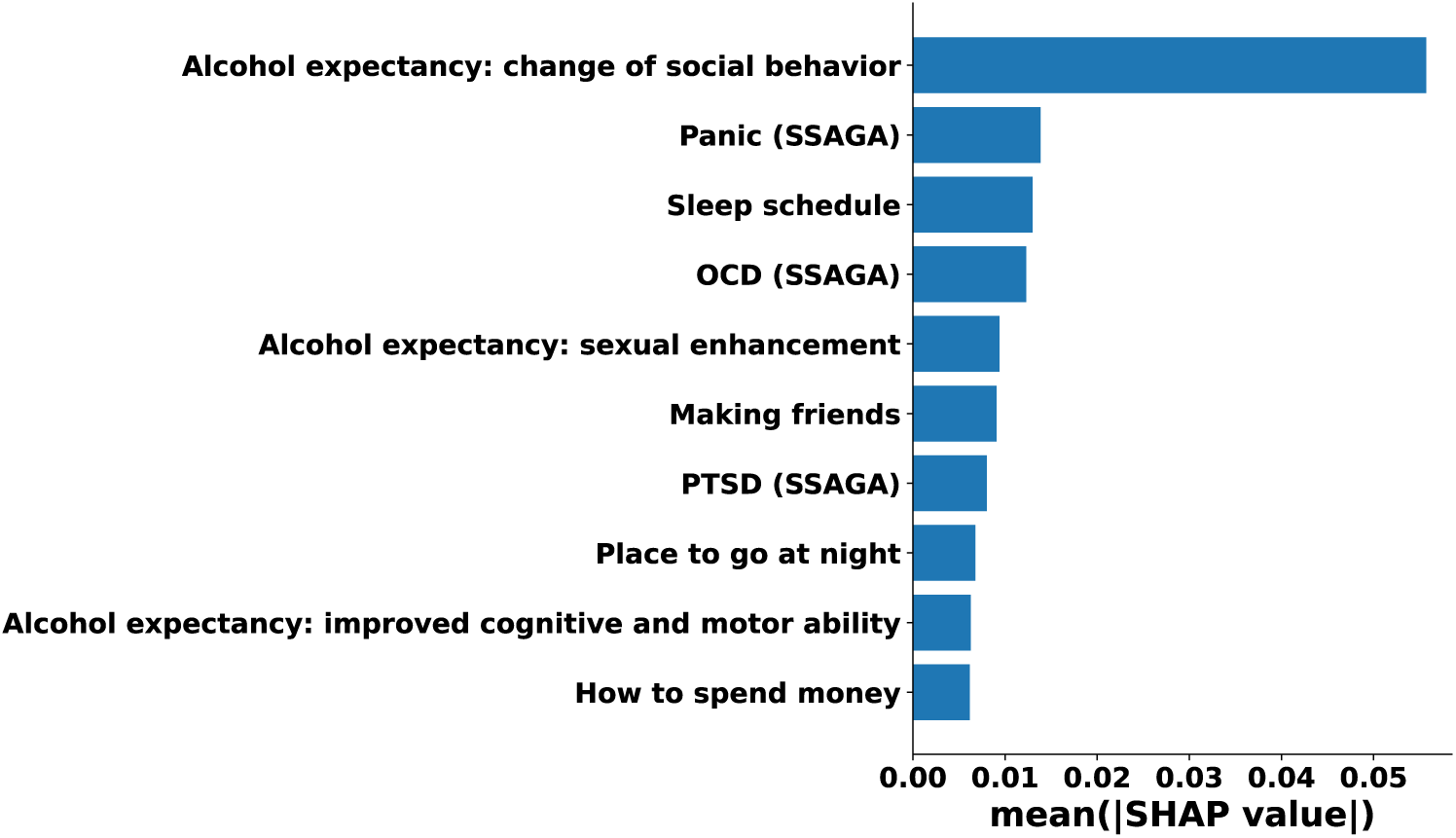
Top 10 most important variables ranked by mean absolute SHAP values from the *FocalTab* model for drinkers classification using the *Imbalanced-Original* dataset with the *w/o Age w/o Substances* variable setting. *Note*: SSAGA refers to Semi-Structured Assessment for the Genetics of Alcoholism.

## 4. Discussion

In this study, we developed a novel TabPFN classification framework with focal loss for distinguishing adolescent drinkers from non-drinkers using exclusively clinical variables obtained through routine medical encounters. Specifically, we utilize a broader adolescent age range (12-22 years), enabling comprehensive modeling across the full developmental trajectory from early adolescence through young adulthood. To mitigate age-related bias, we explicitly account for age confounding through systematic confound regression. In addition, to prevent potential data leakage and inflated predictive performance, substance-use variables are excluded from the feature set. Finally, severe class imbalance is addressed using focal loss rather than synthetic oversampling, thereby preserving the original data distribution while improving the model’s ability to learn from minority-class samples.

The superior performance of *FocalTab* under strict confound control, achieving AUC 0.902, accuracy 0.843, and specificity 0.800 (Table 3), aligns with recent advances in tabular machine learning. TabPFN itself represents a paradigm shift as a foundation model performing in-context learning without parameter updates, achieving normalized AUC 0.939 on the AMLB benchmark and outperforming tuned logistic regression, random forest, and MLP baselines with only 2.8 seconds of compute ^57,63^. Its Bayesian approximation properties and smooth decision boundaries make it particularly suited to clinical tasks with limited samples, and our dataset (N=801) falls within its optimal operating range. When confounds were not fully removed, TabPFN achieved the strongest performance (e.g., AUC 0.912, accuracy 0.770 in the *Balanced w/o Age w/o Substances* setting; Table 5), though combining it with focal loss proved essential under the most stringent conditions.

Our focal loss implementation addressed the substantial class imbalance (661 non-drinkers vs. 140 drinkers; ∼5:1) at the algorithmic level, offering key advantages over data-level approaches such as SMOTE. While competing models on the same imbalanced data exhibited severe majority-class bias, specificity ranging from 0.153 to 0.240 despite sensitivity of 0.807 to 1.000, *FocalTab* maintained a specificity of 0.800 (Table 5). SMOTE yielded only modest specificity gains (e.g., logistic regression 0.200 to 0.393, random forest 0.153 to 0.287) that remained far below *FocalTab*, and actually deteriorated TabPFN’s specificity to 0.107. Focal loss was originally developed for extreme foreground-background imbalance in object detection ^63^, dynamically down-weights well-classified examples to focus learning on hard samples. Prior work confirms its advantage: Baloch et al.^65^ showed that their focal loss variants outperformed both SMOTE and Near-Miss undersampling on 4 out of 6 highly imbalanced fraud detection datasets, achieving gains of up to 3% in balanced accuracy without the computational overhead of synthetic sample generation. In network intrusion detection systems, Mulyanto et al. ^66^ found that while SMOTE combined with deep neural networks achieved acceptable accuracy, F1-scores remained inadequate due to persistent class imbalance issues that focal loss successfully addressed. Beyond performance, focal loss avoids SMOTE’s known drawbacks, including synthetic samples that may misrepresent true distributions, increased class overlap near decision boundaries, and noise amplification in high-dimensional spaces ^40,67^.

Performance across dataset settings followed a consistent ordering: *w/ Age w/ Substances* > *w/ Age w/o Substances* > *w/o Age w/ Substances* > *w/o Age w/o Substances*, indicating that competing models relied heavily on developmental variance and substance use information to achieve apparently high accuracy (Table 4). Specificity declined most sharply upon confound removal: logistic regression fell from 0.760 to 0.200, random forest from 0.767 to 0.153, MLP from 0.753 to 0.173, and TabPFN from 0.787 to 0.167 (Table 4), meaning these models misclassified over 80% of non-drinkers once confounds were controlled. The stronger impact of age removal relative to substance use removal (e.g., random forest AUC 0.911 under *w/ Age w/o Substances* vs. 0.870 under *w/o Age w/ Substances*) reflects the well-documented age-prevalence relationship in adolescent drinking, where prevalence rises from ∼10% at age 14 to over 70% by age 21 ^1^. As Kinreich et al. ^68^ explicitly demonstrated that age is a critical confounding variable in machine learning models for AUD, noting that failure to properly control for age can lead to “misclassification of the model” and biased classifications or predictions. Similarly, the inclusion of other substance use variables (tobacco, marijuana, other drugs) in models introduces potential information leakage, as these substances share common etiological pathways with alcohol use and often co-occur temporally ^69,70^.

Our SHAP ranking of the top 10 most important variables for our model revealed that the most informative variables for distinguishing adolescent drinkers from non-drinkers fell into three clinically meaningful domains (Figure 8): alcohol expectancies, mental health symptoms, and lifestyle characteristics. Alcohol expectancies, which refer to beliefs about the anticipated effects of drinking, emerged as particularly salient variables in our model, with expectancies regarding enhanced social behavior, sexual enhancement, and improved cognitive and motor abilities ranking among the top discriminating features. This is consistent with previous research, demonstrating that positive alcohol expectancies were highly associated with drinking initiation, frequency, and quantity during adolescence ^46,71–74^. Mental health symptoms, including panic, OCD, and PTSD, also emerged as important variables. Epidemiological research has established robust comorbidity between anxiety disorders and AUD, with anxiety disorders being two to three times more prevalent among individuals with alcohol problems than the general population ^75–78^.

This finding is also consistent with the self-medication hypotheses whereby adolescents may use alcohol to manage distressing symptoms ^76,79^. Lifestyle characteristics reflecting adolescents’ daily routines and social environments constituted the third major classification domain, encompassing sleep patterns, friend-making, nighttime activities, and spending money behaviors. Sleep disturbances have been increasingly recognized as both risk factors for and consequences of adolescent alcohol use^80^. Friend-making is consistent with the extensive literature documenting peer friends’ influence as one of the most proximal risk factors for adolescent substance use ^81,82^. Unstructured, unsupervised socializing with peers creates opportunities for risk-taking behavior, including substance use ^83,84^. Finally, adolescent ways of spending money, relating to disposable income, have been identified as a significant predictor of drinking behavior ^85^ - adolescents with a higher level of money spending have greater access to alcohol and may participate in social contexts where drinking is more prevalent ^86^.

Several limitations should be acknowledged. First, although the sample size of 801 participants was adequate for the current analyses, it may limit the generalizability of our findings to more diverse adolescent populations. Second, formal external validation using an independent dataset was not conducted. Future work will address these limitations by validating the proposed framework in larger and independent cohorts. In addition, we will extend our work to longitudinal analyses to prospectively predict drinking status across subsequent NCANDA follow-up assessments.

## Data Availability

All data produced in the present study are available upon reasonable request to the authors

## Acknowledgment

Research reported in this publication was supported by the U.S. National Science Foundation under Award Number 2500836, and the Office of the Director, National Institutes of Health of the National Institutes of Health under Award Number R03OD038391. This work was also partially supported by the National Institute of General Medical Sciences (NIGMS) under Award Numbers P20GM103427 and U54GM115458, and by the National Institute on Alcohol Abuse and Alcoholism (NIAAA) under Award Numbers R01 AA029127, P60 AA031124, F32 AA032170, and L30 AA032656, P50AA030407-5126 (Pilot Core grant). This study was in part financially supported by the Child Health Research Institute at UNMC/Children’s Nebraska. This work was also partially supported by Nebraska EPSCoR FIRST Award (OIA-2044049). The content is solely the responsibility of the authors and does not necessarily represent the official views of the funding organizations.

## References

1. Substance Abuse and Mental Health Services Administration (SAMHSA), Center for Behavioral Health Statistics and Quality (CBHSQ). 2024 National Survey on Drug Use and Health. Table 2.44A—Alcohol use in lifetime, past year, and past month and binge alcohol and heavy alcohol use in past month: among people aged 12 to 20; by demographic characteristics, numbers in thousands, 2023 and 2024. (2024).

2. Niaaa, P.-T. D. A., Navigator, A. T. & Plan, N. S. Alcohol screening and brief intervention for youth: a practitioner’s guide. (2011).

3. Boden, J. M. & Fergusson, D. M. The short-and long-term consequences of adolescent alcohol use. Young people and alcohol: Impact, policy, prevention, treatment 32–44 (2011).

4. Marshall, E. J. Adolescent alcohol use: risks and consequences. Alcohol and alcoholism 49, 160–164 (2014).

5. Brassiolo, P., Fagre, E., Harker Roa, A. & Peñaloza-Pacheco, L. Preventing underage alcohol use through changes in norms and risk perception: A randomized evaluation of two school-based prevention programs in Colombia. (2023).

6. Esposito, S. et al. Too young to pour: the global crisis of underage alcohol use. Frontiers in public health 13, 1598175 (2025).

7. Tanner-Smith, E. E. & Lipsey, M. W. Brief alcohol interventions for adolescents and young adults: A systematic review and meta-analysis. Journal of substance abuse treatment 51, 1–18 (2015).

8. Zhu, T. et al. Machine learning of functional connectivity to biotype alcohol and nicotine use disorders. Biological Psychiatry: Cognitive Neuroscience and Neuroimaging 9, 326–336 (2024).

9. Vergara, V. M., Espinoza, F. A. & Calhoun, V. D. Identifying alcohol use disorder with resting state functional Magnetic Resonance Imaging data: a comparison among machine learning classifiers. Frontiers in Psychology 13, 867067 (2022).

10. Zhu, X., Du, X., Kerich, M., Lohoff, F. W. & Momenan, R. Random forest based classification of alcohol dependence patients and healthy controls using resting state MRI. Neurosci Lett 676, 27–33 (2018).

11. Whelan, R. et al. Neuropsychosocial profiles of current and future adolescent alcohol misusers. Nature 512, 185–189 (2014).

12. Ottino-González, J. et al. Brain structural covariance network features are robust markers of early heavy alcohol use. Addiction 119, 113–124 (2024).

13. Afzali, M. H. et al. Machine-learning prediction of adolescent alcohol use: a cross-study, cross-cultural validation. Addiction 114, 662–671 (2019).

14. Ebrahimi, A., Wiil, U. K., Andersen, K., Mansourvar, M. & Nielsen, A. S. A predictive machine learning model to determine alcohol use disorder. in 2020 IEEE Symposium on Computers and Communications (ISCC) 1-7 (IEEE, 2020).

15. Lee, M. R. et al. Using Machine Learning to Classify Individuals With Alcohol Use Disorder Based on Treatment Seeking Status. EClinicalMedicine 12, 70–78 (2019).

16. Hahn, S. et al. Predicting alcohol dependence from multi-site brain structural measures. Human Brain Mapping 43, 555–565 (2022).

17. Lee, S. Development of deep learning auto-encoder algorithms for predicting alcohol use in Korean adolescents based on cross-sectional data. Social Science & Medicine 367, 117690 (2025).

18. Englund, M. M., Egeland, B., Oliva, E. M. & Collins, W. A. Childhood and adolescent predictors of heavy drinking and alcohol use disorders in early adulthood: a longitudinal developmental analysis. Addiction 103, 23–35 (2008).

19. Pedroni, C. et al. Alcohol consumption in early adolescence: Associations with sociodemographic and psychosocial factors according to gender. PLoS One 16, e0245597 (2021).

20. Carriedo, N., Rodríguez-Villagra, O. A., Moriano, J. A., Montoro, P. R. & Iglesias-Sarmiento, V. Age-related change in inhibitory processes when controlling working memory capacity and processing speed: A confirmatory factor analysis. Plos one 20, e0316347 (2025).

21. Ferguson, H. J., Brunsdon, V. E. & Bradford, E. E. The developmental trajectories of executive function from adolescence to old age. Scientific reports 11, 1382 (2021).

22. Skalaban, L. J. et al. Adolescent-specific memory effects: evidence from working memory, immediate and long-term recognition memory performance in 8-30 yr olds. Learning & Memory 29, 223–233 (2022).

23. Steinberg, L. et al. Age differences in sensation seeking and impulsivity as indexed by behavior and self-report: evidence for a dual systems model. Developmental Psychology 44, 1764–1778 (2008).

24. Rogers, A. A., Padilla-Walker, L. M., McLean, R. D. & Hurst, J. L. Trajectories of perceived parental psychological control across adolescence and implications for the development of depressive and anxiety symptoms. Journal of youth and adolescence 49, 136–149 (2020).

25. Laursen, B. & Veenstra, R. Toward understanding the functions of peer influence: A summary and synthesis of recent empirical research. Journal of Research on Adolescence 31, 889–907 (2021).

26. Dell, N. A. et al. Binge drinking in early adulthood: A machine learning approach. Addictive Behaviors 124, 107122 (2022).

27. Tschorn, M. et al. Differential predictors for alcohol use in adolescents as a function of familial risk. Translational psychiatry 11, 157 (2021).

28. Trucco, E. M. A review of psychosocial factors linked to adolescent substance use. Pharmacology biochemistry and behavior 196, 172969 (2020).

29. National Institutes of Health. Reported drug use among adolescents continued to hold below pre-pandemic levels in 2023. (2023).

30. Abuse, S. & others. Key substance use and mental health indicators in the United States: results from the 2019 National Survey on Drug Use and Health. (2020).

31. U.S. Food & Drug Administration. Youth Tobacco Use: Results from the National Youth Tobacco Survey. (2020).

32. Wang, T. W. E-cigarette use among middle and high school students—United States, 2020. MMWR. Morbidity and Mortality Weekly Report 69, (2020).

33. Park-Lee, E. Notes from the field: e-cigarette use among middle and high school students—National Youth Tobacco Survey, United States, 2021. MMWR. Morbidity and Mortality Weekly Report 70, (2021).

34. Wang, T. W. Tobacco product use and associated factors among middle and high school students—United States, 2019. MMWR. Surveillance Summaries 68, (2019).

35. Gentzke, A. S. Tobacco product use among middle and high school students—United States, 2020. MMWR. Morbidity and Mortality Weekly Report 69, (2020).

36. Yang, J., Mejia, M. C., Sacca, L., Hennekens, C. H. & Kitsantas, P. Trends in marijuana use among adolescents in the United States. Pediatric Reports 16, 872–879 (2024).

37. Harris, K. M., Griffin, B. A., McCaffrey, D. F. & Morral, A. R. Inconsistencies in self-reported drug use by adolescents in substance abuse treatment: implications for outcome and performance measurements. Journal of Substance Abuse Treatment 34, 347–355 (2008).

38. Khalili, P. et al. Validity of self-reported substance use: research setting versus primary health care setting. Substance abuse treatment, prevention, and policy 16, 66 (2021).

39. Wade, N. E., Patel, H. & Pelham III, W. E. A comparison of remote versus in-person assessments of substance use and related constructs among adolescents. Substance use & misuse 59, 1447–1454 (2024).

40. Chawla, N. V., Bowyer, K. W., Hall, L. O. & Kegelmeyer, W. P. SMOTE: synthetic minority over-sampling technique. Journal of artificial intelligence research 16, 321–357 (2002).

41. Zhao, Q. et al. Identifying high school risk factors that forecast heavy drinking onset in understudied young adults. Developmental Cognitive Neuroscience 68, 101413 (2024).

42. Lin, T.-Y., Goyal, P., Girshick, R., He, K. & Dollár, P. Focal loss for dense object detection. in Proceedings of the IEEE international conference on computer vision 2980–2988 (2017).

43. Brown, S. A. et al. The National Consortium on Alcohol and NeuroDevelopment in Adolescence (NCANDA): a multisite study of adolescent development and substance use. Journal of studies on alcohol and drugs 76, 895–908 (2015).

44. Cahalan, D., Cisin, I. H. & Crossley, H. M. American Drinking Practices: A National Study of Drinking Behavior and Attitudes. (Rutgers Center of Alcohol Studies, 1969).

45. Brown, S. A. et al. Psychometric evaluation of the Customary Drinking and Drug Use Record (CDDR): a measure of adolescent alcohol and drug involvement. Journal of studies on alcohol 59, 427–438 (1998).

46. Brown, S. A., Christiansen, B. A. & Goldman, M. S. The Alcohol Expectancy Questionnaire: an instrument for the assessment of adolescent and adult alcohol expectancies. Journal of studies on alcohol 48, 483–491 (1987).

47. Rice, J. P. et al. Comparison of direct interview and family history diagnoses of alcohol dependence. Alcoholism: Clinical and Experimental Research 19, 1018–1023 (1995).

48. Hollingshead, A. B. Four Factor Index of Social Status. (1975).

49. Gosling, S. D., Rentfrow, P. J. & Swann Jr, W. B. A very brief measure of the Big-Five personality domains. Journal of Research in personality 37, 504–528 (2003).

50. Cyders, M. A. et al. Integration of impulsivity and positive mood to predict risky behavior: development and validation of a measure of positive urgency. Psychological assessment 19, 107 (2007).

51. Gioia, G. A., Isquith, P. K., Retzlaff, P. D. & Espy, K. A. Confirmatory factor analysis of the Behavior Rating Inventory of Executive Function (BRIEF) in a clinical sample. Child neuropsychology 8, 249–257 (2002).

52. Connor-Smith, J. K., Compas, B. E., Wadsworth, M. E., Thomsen, A. H. & Saltzman, H. Responses to stress in adolescence: measurement of coping and involuntary stress responses. Journal of consulting and clinical psychology 68, 976 (2000).

53. Petersen, A. C., Crockett, L., Richards, M. & Boxer, A. A self-report measure of pubertal status: Reliability, validity, and initial norms. Journal of youth and adolescence 17, 117–133 (1988).

54. Sarason, I. G., Sarason, B. R., Shearin, E. N. & Pierce, G. R. A brief measure of social support: Practical and theoretical implications. Journal of social and personal relationships 4, 497–510 (1987).

55. Bucholz, K. K. et al. A new, semi-structured psychiatric interview for use in genetic linkage studies: a report on the reliability of the SSAGA. Journal of studies on alcohol 55, 149–158 (1994).

56. Radloff, L. S. The CES-D scale: A self-report depression scale for research in the general population. Applied psychological measurement 1, 385–401 (1977).

57. Hollmann, N., Müller, S., Eggensperger, K. & Hutter, F. Tabpfn: A transformer that solves small tabular classification problems in a second. arXiv preprint arXiv:2207.01848 (2022).

58. LaValley, M. P. Logistic regression. Circulation 117, 2395–2399 (2008).

59. Rigatti, S. J. Random forest. Journal of insurance medicine 47, 31–39 (2017).

60. Ruck, D. W., Rogers, S. K. & Kabrisky, M. Feature selection using a multilayer perceptron. Journal of neural network computing 2, 40–48 (1990).

61. Mannor, S., Peleg, D. & Rubinstein, R. The cross entropy method for classification. In Proceedings of the 22nd international conference on Machine learning 561–568 (2005).

62. Lundberg, S. M. & Lee, S.-I. A unified approach to interpreting model predictions. Advances in neural information processing systems 30, (2017).

63. Hollmann, N. et al. Accurate predictions on small data with a tabular foundation model. Nature 637, 319–326 (2025).

64. Lin, T.-Y., Goyal, P., Girshick, R., He, K. & Dollár, P. Focal loss for dense object detection. in Proceedings of the IEEE international conference on computer vision 2980-2988 (2017).

65. Baloch, B. K., Kumar, S., Haresh, S., Rehman, A. & Syed, T. Focused Anchors Loss: cost-sensitive learning of discriminative features for imbalanced classification. in Proceedings of The Eleventh Asian Conference on Machine Learning vol. 101 822–835 (2019).

66. Mulyanto, M., Faisal, M., Prakosa, S. W. & Leu, J.-S. Effectiveness of Focal Loss for Minority Classification in Network Intrusion Detection Systems. Symmetry 13, 4 (2021).

67. Cui, Y., Jia, M., Lin, T.-Y., Song, Y. & Belongie, S. Class-Balanced Loss Based on Effective Number of Samples. in 2019 IEEE/CVF Conference on Computer Vision and Pattern Recognition (CVPR) 9260–9269 (2019). doi:10.1109/CVPR.2019.00949.

68. Kinreich, S. et al. Predicting risk for Alcohol Use Disorder using longitudinal data with multimodal biomarkers and family history: a machine learning study. Molecular Psychiatry 26, 1133–1141 (2021).

69. Brown, S. A. et al. A developmental perspective on alcohol and youths 16 to 20 years of age. Pediatrics 121, S290–S310 (2008).

70. Chassin, L. et al. Adolescent substance use. in Handbook of Adolescent Psychology (eds Lerner, R. M. & Steinberg, L.) 665–696 (John Wiley & Sons, Inc., 2004). doi:10.1002/9780471726746.ch21.

71. Christiansen, B. A., Smith, G. T., Roehling, P. V. & Goldman, M. S. Using alcohol expectancies to predict adolescent drinking behavior after one year. Journal of Consulting and Clinical Psychology 57, 93–99 (1989).

72. Copeland, A. L., Proctor, S. L., Terlecki, M. A., Kulesza, M. & Williamson, D. A. Do positive alcohol expectancies have a critical developmental period in pre-adolescents? Journal of Studies on Alcohol and Drugs 75, 945–952 (2014).

73. Smith, G. T., Goldman, M. S., Greenbaum, P. E. & Christiansen, B. A. Expectancy for social facilitation from drinking: the divergent paths of high-expectancy and low-expectancy adolescents. Journal of Abnormal Psychology 104, 32–40 (1995).

74. Stein, L. A. et al. Validity and Reliability of the Alcohol Expectancy Questionnaire-Adolescent, Brief. Journal of Child & Adolescent Substance Abuse 16, 115–127 (2007).

75. Back, S. E. & Brady, K. T. Anxiety Disorders with Comorbid Substance Use Disorders: Diagnostic and Treatment Considerations. Psychiatric Annals 38, 724–729 (2008).

76. Buckner, J. D. et al. Specificity of social anxiety disorder as a risk factor for alcohol and cannabis dependence. Journal of Psychiatric Research 42, 230–239 (2008).

77. Kessler, R. C., Chiu, W. T., Demler, O., Merikangas, K. R. & Walters, E. E. Prevalence, severity, and comorbidity of 12-month DSM-IV disorders in the National Comorbidity Survey Replication. Archives of General Psychiatry 62, 617–627 (2005).

78. Smith, J. P. & Book, S. W. Anxiety and Substance Use Disorders: A Review. Psychiatric Times 25, 19–23 (2008).

79. Wolitzky-Taylor, K., Bobova, L., Zinbarg, R. E., Mineka, S. & Craske, M. G. Longitudinal investigation of the impact of anxiety and mood disorders in adolescence on subsequent substance use disorder onset and vice versa. Addictive Behaviors 37, 982–985 (2012).

80. Hasler, B. P., Soehner, A. M. & Clark, D. B. Sleep and circadian contributions to adolescent alcohol use disorder. Alcohol 49, 377–387 (2015).

81. Ali, M. M. & Dwyer, D. S. Social network effects in alcohol consumption among adolescents. Addictive Behaviors 35, 337–342 (2010).

82. Brechwald, W. A. & Prinstein, M. J. Beyond Homophily: A Decade of Advances in Understanding Peer Influence Processes. Journal of Research on Adolescence 21, 166–179 (2011).

83. Lee, K. T. H. & Vandell, D. L. Out-of-School Time and Adolescent Substance Use. Journal of Adolescent Health 57, 523–529 (2015).

84. Osgood, D. W., Anderson, A. L. & Shaffer, J. N. Unstructured leisure in the after-school hours. in Organized Activities as Contexts of Development: Extracurricular Activities, After-School and Community Programs (eds Mahoney, J. L., Larson, R. W. & Eccles, J. S.) 45–64 (Lawrence Erlbaum Associates Publishers, 2005).

85. Brunborg, G. S., von Soest, T. & Burdzovic Andreas, J. Adolescent income and binge drinking initiation: prospective evidence from the MyLife study. Addiction 116, 1389–1398 (2021).

86. Lintonen, T. & Nevalainen, J. Has the role of personal income in alcohol drinking among teenagers changed between 1983 and 2013: a series of nationally representative surveys in Finland. BMJ Open 7, e013994 (2017).

